# Developing Consensus for an Upper and Lower Limb Athlete Pain Assessment Framework – A Real-time Delphi Study with International Sports Physiotherapists

**DOI:** 10.1101/2024.06.14.24308931

**Authors:** Ciarán Purcell, Brona M Fullen, Tomás Ward, Brian M Caulfield

## Abstract

**Objectives:** There is no current consensus on the key items sports physiotherapists should consider when completing a comprehensive biopsychosocial upper or lower limb pain assessment with athletes. We sought to develop recommendations to inform a framework for the assessment of upper and lower limb pain in athletes.

**Design:** Real-time Delphi

**Methods:** We recruited sports physiotherapists currently working with athletes through the International Society of Sports Physical Therapists and Irish Society of Chartered Physiotherapists. Participants voted on 86 pain assessment items chosen using best available evidence. The real-time Delphi method facilitated independent anonymous voting, commenting and immediate review of consensus. Participants indicated level of agreement for inclusion in an upper and lower limb athlete pain assessment framework on a 6-point Likert scale from Strongly Disagree to Strongly Agree, and how often they are/will be required in practice on a 5-point scale from *Never* to *Always.* Criteria for consensus agreement and inclusion were i) >70% sports physiotherapists voting agree/strongly agree AND ii) median vote selected by physiotherapists was Agree or Strongly Agree.

**Results:** 41 sports physiotherapists (female n=20, male n=21), visited the survey an average of 5.3 times (±5), resulting in a completion rate of 98%. 64 assessment items (neurophysiological n=20, biomechanical n=15, affective n=8, cognitive n=3, socioenvironmental n=10, general assessment aspects of assessment n=8) met the criteria for consensus. Frequency of use in practice was *Always* for 28 items *Often* for 32 items and *Sometimes* for 4 items.

**Conclusion:** We have presented stakeholder-generated recommendations and priorities for athlete pain assessment.

## Introduction

Pain is a complex biopsychosocial experience, with sensory and affective aspects.^44^ The modern predictive processing model sheds light on how pain shapes the way we interact in the world which both influences and is influenced by a host of internal and external contextual and environmental factors.^29^ Pain assessment includes embracing the multidimensional nature of pain which has led to pain assessment guidelines for many aspects of musculoskeletal pain.^13, 30^ Pain in athletes is a lesser- explored phenomenon that significantly impacts performance and well-being yet is often undervalued due in part to a focus on a time-loss definition of injury severity reporting in research and a “play through pain” sports culture that prioritises availability to compete.^1, 3^ The subjective, individual and complex nature of pain poses a challenge for time-pressured clinicians and athletes.^28, 39^

The International Olympic Committee’s (IOC) consensus statement on pain in elite athletes and related works recommend a comprehensive multidimensional assessment of pain.^22, 23, 26^ A scoping review of the assessment tools for athletes experiencing pain in the upper or lower limb identified 175 tools used in research and practice, 77 of which were used in nine or more studies.^42^ The review in the IOC consensus (neurophysiological n =32, biomechanical n =96, affective n=21, cognitive n=8, socioenvironmental n=18).^22^ A gap in the use of tools assessing the the internal contextual factors (affective and cognitive) and external contextual factors (socioenvironmental) was highlighted, aligning with gaps identified by the IOC.^57^ Qualitative research harnessing the athlete’s voice has is an effective method to explore the comprehensive nature of pain in athletes, with insights identifying the importance of contextual factors when it comes to understanding pain and making informed decisions.^2, 6, 10^ A series of focus groups of athletes and sports physiotherapists^40, 41^ highlighted a need for increased integration of multidimensional pain assessment methods alongside considering aspects surrounding the assessment process such as clear and timely communication and developing strong therapeutic relationships, aligning with contemporary athlete pain research.^8, 25^

Despite research highlighting a comprehensive list of important components for a multidimensional assessment that athletes and sports physiotherapists *could* consider, there is no consensus on how they *should* be applied. A practical, clinician-friendly, integrative framework that identifies the most important and relevant pain assessment tools for use across a range of athletic cohorts is lacking. It is unclear how and when a sports physiotherapist should apply the available tools when completing an upper or lower limb pain assessment with an athlete. There is a need to develop an upper and lower limb pain assessment framework that is comprehensive, effectively representing an athlete’s pain experience, whilst also being practical to facilitate implementation in clinical practice.

We aimed to generate sports physiotherapist consensus-derived recommendations for an upper and lower limb athlete pain assessment framework for research and clinical practice using the real-time Delphi (RTD) consensus method.

## Methods

This RTD study followed contemporary reporting guidelines developed for Delphi techniques in the health sciences and the ACcurate COnsensus Reporting Document (ACCORD) reporting guidelines.^17, 51^ Ethical permission was granted for our study by the Universities’ Human Research Ethics Committee. (LS-22-40-xxx-xxx) Informed consent was provided by each participant and the rights of participants were protected throughout the study.

### Participants

An international panel of sports physiotherapists was established through purposive sampling and recruitment via the Irish Society of Chartered Physiotherapists Sports and Exercise Medicine Special Interest Group and the International Federation of Sports Physical Therapy. A Group of 30-50 Sports Physiotherapists, with three years minimum postgraduate experience working with athletes as part of their weekly caseload were sought, allowing for representation in the breadth of experience, gender, geographical location, sports caseload and assessment perspectives whilst also facilitating consensus development.^14, 46, 54^

### Patient, athletes and public involvement

Five athletes, (n=3 female, n=2 male) across a range of competition levels and sports formed a PPI panel. They initially reviewed and commented on the research questions and initial survey and their feedback was incorporated into the design and delivery of the RTD. The athletes were subsequently consulted regarding the analysis and presentation of the results.

### Protocol

The Delphi technique is a method of consensus generation suited to exploring complex phenomena including pain.^17^ It facilitates the equal contribution of participants and identifies areas of convergence as well as divergence providing valuable insights.^51^ The RTD method facilitates participants to view evolving consensus in real-time, contribute comments and revisit their responses and consensus multiple times throughout the study without the burden of engagement and analysis required in traditional round-based Delphi studies.^19, 20^ The RTD format has been validated against traditional approaches with researchers commenting on its practicality and efficiency within healthcare settings.^37, 43^

Using the Surveylet commercial software package (Calibrum Inc, Utah, USA) sports physiotherapists anonymously completed baseline demographics followed by the RTD. The initial survey (Appendix A) contained 86 assessment items across six domains (neurophysiological, biomechanical, affective, cognitive, socioenvironmental and general aspects of assessment). The survey was designed using the IOC athlete pain consensus statement and related works,^22, 23^ findings from a scoping review of athlete pain assessments,^42^ focus groups exploring athlete pain assessment priorities from shared athlete and physiotherapist perspectives,^40, 41^ and additional qualitative research.^2, 10^ Feedback from two expert physiotherapist pain researchers, with over 25 years of experience was sought alongside consultation with the athlete PPI panel. Following an initial pilot with three members of the research team, the RTD survey was released to all participants on November 10^th^, 2023, and ran for eight weeks. Weekly reminders were sent to all participants yet to complete the survey and fortnightly emails were sent to encourage participants to revisit the survey, review the evolving consensus and contribute comments as often as they wished, facilitating engagement.

### Consensus

The survey was designed to address; i) whether participants felt each assessment items *should* be included in an athlete upper and lower limb pain assessment framework, as well as ii) how often they would expect these assessment items to be required. Participants were asked to indicate their level of agreement for (i) using a 6-point Likert scale (strongly disagree–disagree–-somewhat disagree– somewhat agree–agree– strongly agree) which was chosen as a valid measure to establish consensus. It has been used in previous research to generate expert physiotherapist consensus and avoids ambivalent participant voting.^46^ Consensus agreement criteria was defined a priori; i) a minimum of 70% of participants either *agree* or *strongly agree* with the inclusion of the item. AND ii) the median vote must be either *agree* or *strongly agree* for the item to meet consensus threshold for inclusion. iii) Items where a minimum of 70% of participants voted either *strongly disagree* or *disagree* met the consensus threshold for exclusion. iv) All other items would therefore fail to reach the threshold for consensus. We chose a consensus threshold of 70% in line with consensus agreement guidelines that discuss the value of setting a specific threshold (51-90%) for a specific portion of votes within a certain range alongside the median value.^15^ In this case we chose a relatively narrow range (2 points on the 6 point Likert scale, Agree and Strongly Agree). 70% was chosen to facilitate consensus on important items whilst ensuring sufficient agreement and has been used in similar studies seeking consensus from Physiotherapists.^46^

Participants were encouraged to think about the potential practicality and feasibility of the pain assessment items and asked to select how often they felt each of the pain assessment items are/will be required in practice on a 5-point scale. (*never – rarely – sometimes – often – always* The mean score was reported. Participants were able to view the current consensus immediately upon completion of their initial voting. (Appendix B) The number of times participants reviewed the evolving consensus for each item, changed their score and added a comment on an assessment item are reported in the results.

## Results

### Respondents

In total, 49 sports physiotherapists responded to our invitation. Of those, 41 sports physiotherapists (N=20 female, N= 21 male) took part in the RTD. 38 participants completed the entire survey (100%). Three participants completed 90%, 65% and 55% of the survey respectively. Of the 41 participants who began the survey, the mean completion rate was 98 %. Those who responded to the initial survey but did not complete any of the survey sections were removed from analysis.

Sports physiotherapists were from a number of countries (Ireland n=20, UK n=7, US n=4, South Africa n=4, Spain n=2, Australia n=2, Sweden n=1, Switzerland n=1) and worked with athletes from a range of sports (skill, power, endurance and mixed).^36^ Participants had a mean of 15 years (+/- 5.7) post-graduate experience with 28 participants (68%) having completed additional sports physiotherapy/sports medicine postgraduate education and 33 participants (80%) having sports physiotherapist accreditation. Participants visited the survey an average of 5.3 times (±5, median - 4). See Figure 1 for the full participant descriptive statistics.

**FIGURE 1.**
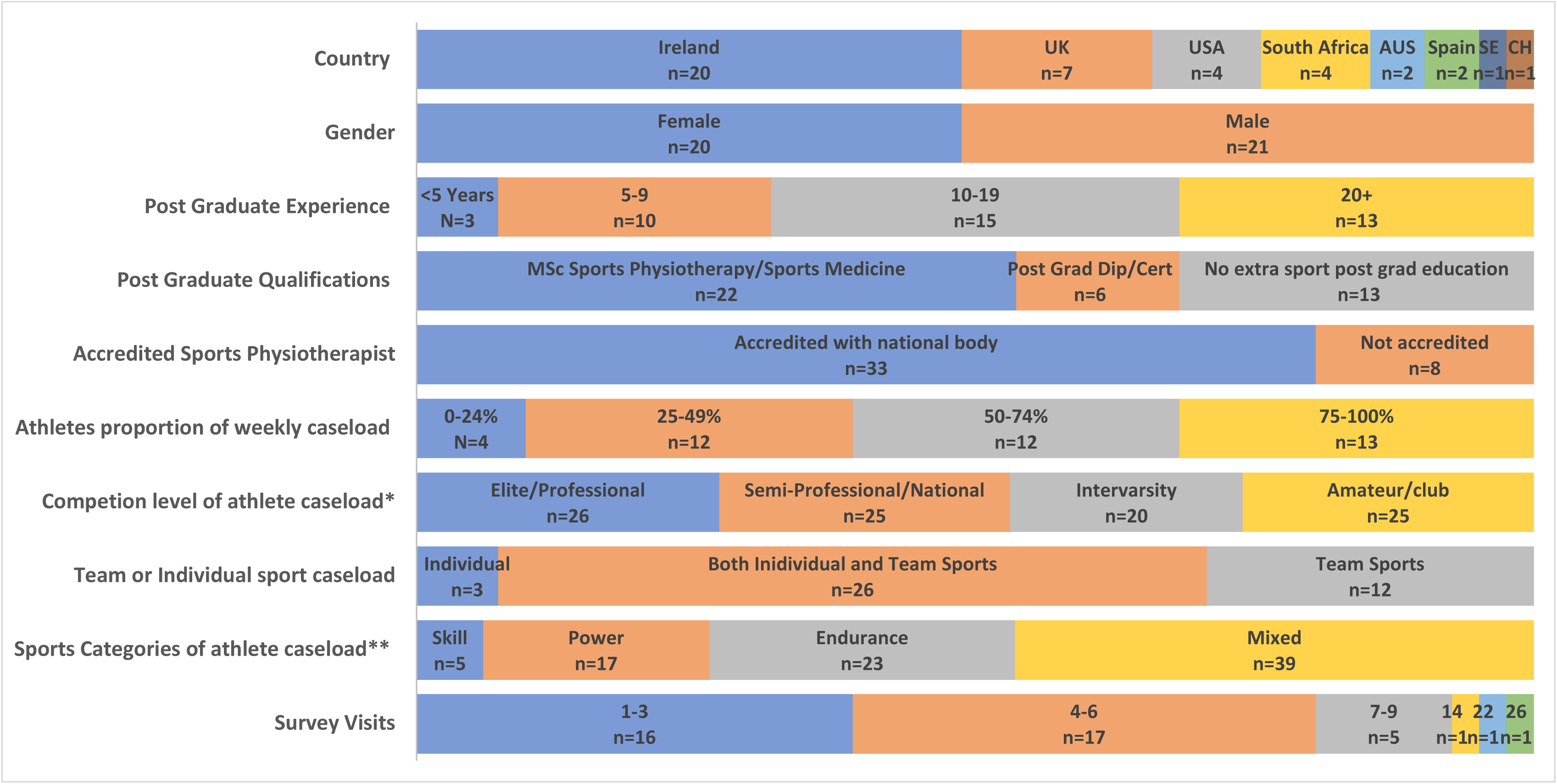
Participant Demographics.

### Response and engagement

Consensus for each assessment item was viewed on average 98 times (+/-31, median=105). The RTD presented the assessment items for each of the six domains on a separate page. Therefore, consensus was viewed an equal number of times for all items in each domain. Consensus was viewed by all participants for all items at least once. Pain assessment items in the neurophysiological domain were reviewed on average 2.6 times (+/- 1.6, median=2) per participant. The biomechanical items were reviewed 3.6 (+/- 4.5, median=2) times, the affective items 2 (+/-1.9, median=1) times, the cognitive items 1.7 (+/-1.3, median=1) times the socioenvironmental items 1.6 (+/- 1.3. median=1) times and the general aspects of pain assessment items 1.9 (+/- 1.7, median =1) times.

Of the 8472 consensus reviews, there were 66 participant changes in consensus vote. The mean change in consensus vote was 1.3 (+/-0.7) on the 6-point Likert scale. There was a total of 218 comments made by 18 participants across the 86 pain assessment items. There was a mean of 2.2 (+/- 1.3) comments on items that achieved consensus and 3.5 (+/- 2.1) comments on items that did not.

### Section 1 - Neurophysiological Domain

Table 1 displays the level of consensus reached for each of the neurophysiological pain assessment items. 20 out of 27 neurophysiological items met the consensus threshold for inclusion. Participants indicated that 16 neurophysiological items are *always* and four are *often* required in practice.

**TABLE 1.**
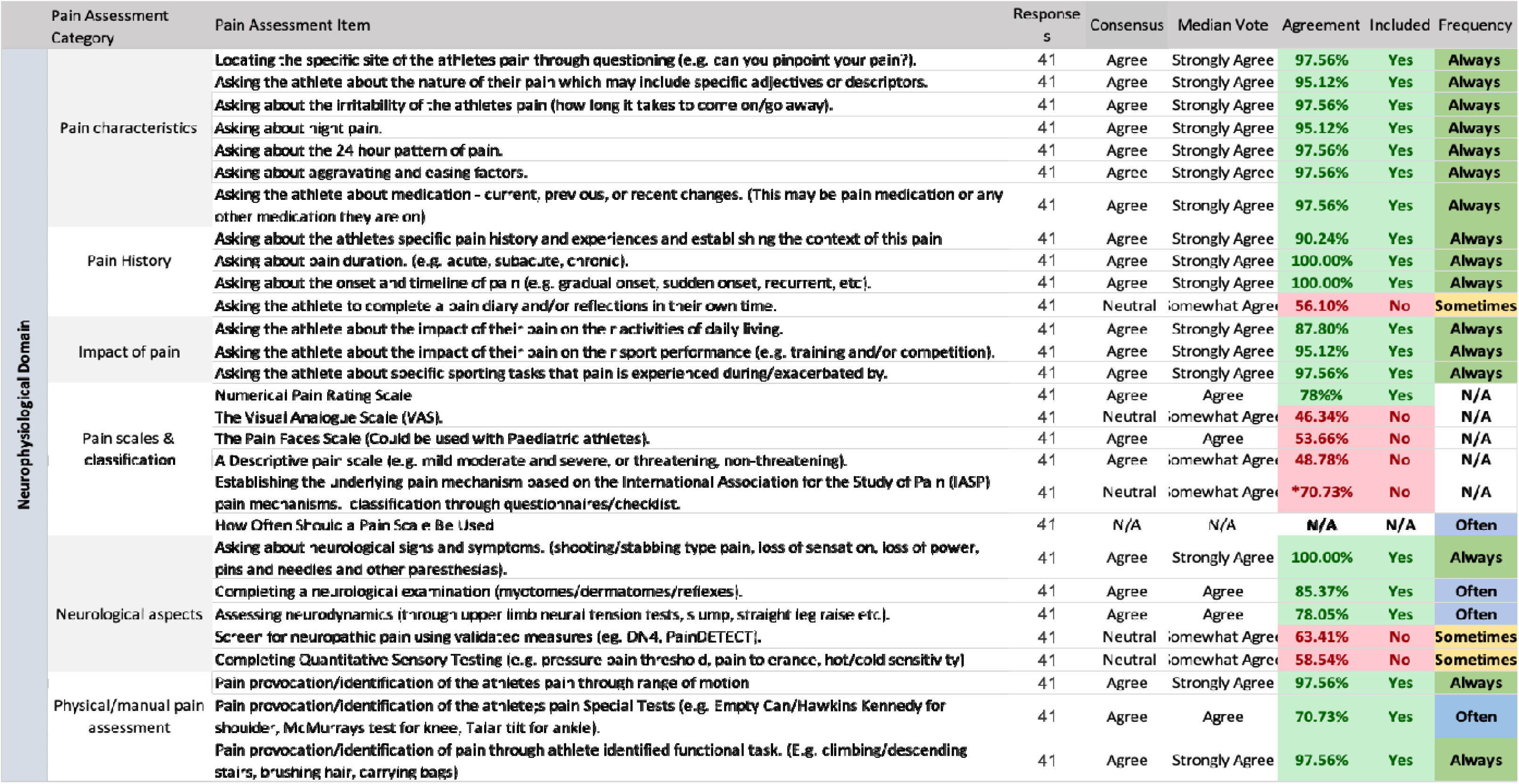
Neurophysiological Domain Assessment Items and Consensus. *items where >70% of participants voted either somewhat agree or somewhat disagree indicating a neutral consensus

### Section 2 - Biomechanical and Affective Domains

Table 2 displays the level of consensus reached for each of the biomechanical and affective pain assessment items. 15 out of 20 biomechanical items met the consensus threshold for inclusion: six *always*, nine *often.* Participants voted to include eight out of nine affective items: seven *often* and one *sometimes*.

**TABLE 2.**
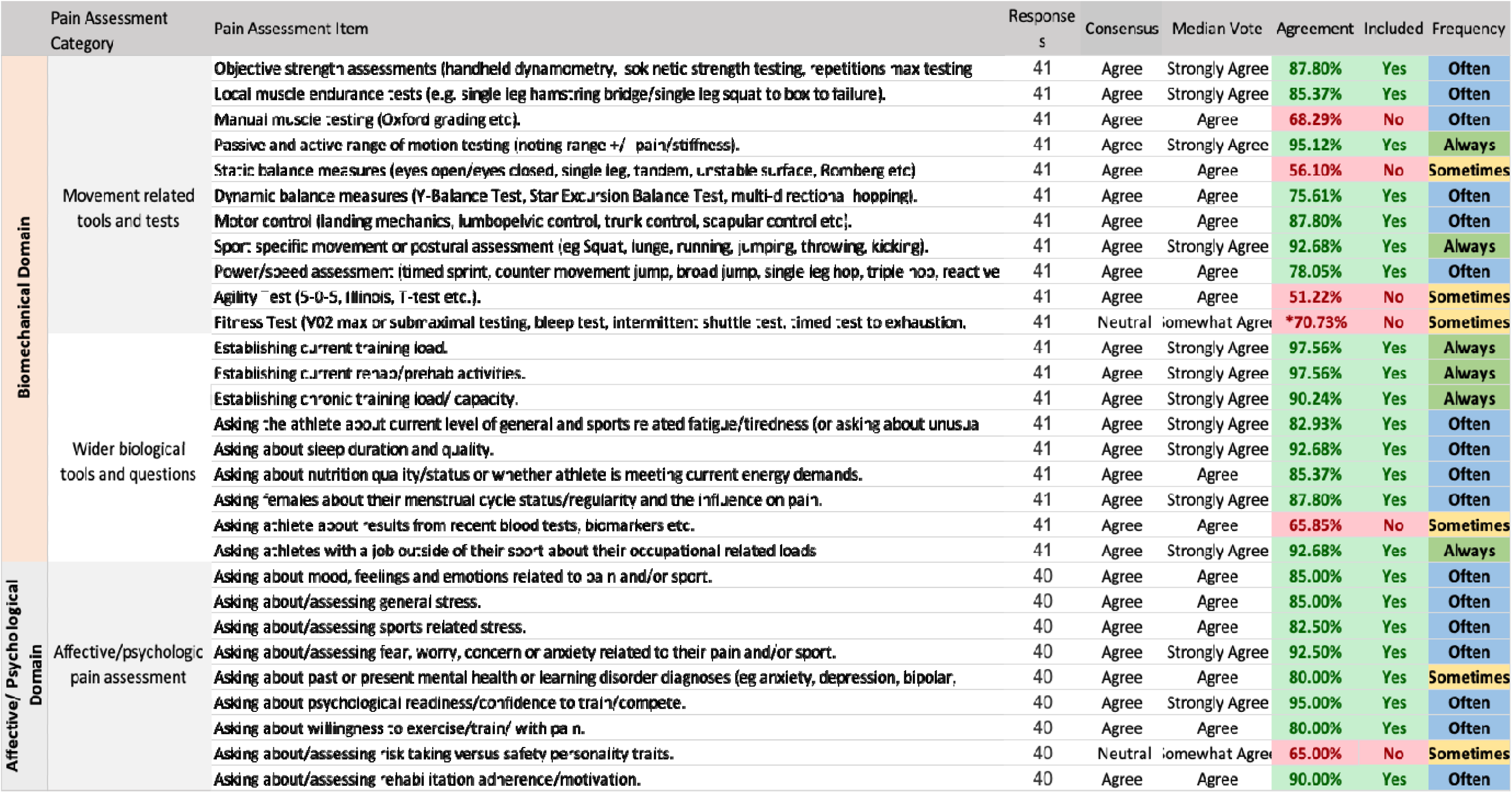
Biomechanical and Affective Domain Assessment Items and Consensus. *items where >70% of participants voted either somewhat agree or somewhat disagree indicating a neutral consensus

### Section 3 – Cognitive, Socioenvironmental and General Aspects of Assessment Domains

Table 3 displays the level of consensus reached for each of the cognitive, socioenvironmental and general aspects of assessment items. Three out of six cognitive domain items met the consensus threshold for inclusion, participants indicated all three are required *often.* Participants voted to include ten out of 15 socioenvironmental items: seven *often* and three *sometimes.* Eight out of nine general aspects of assessment met the consensus threshold for inclusion: six *always*, two *often*. In total, 64 items met consensus for inclusion, no items met consensus for exclusion. 22 items failed to meet consensus.

**TABLE 3.** Cognitive, Socioenvironmental and General Aspects of Assessment Domain Items and Consensu.

|  | Pain Assessment Category | Pain Assessment Item | Responses | Consensus | Median Vote | Agreement | Included | Frequency |
| --- | --- | --- | --- | --- | --- | --- | --- | --- |
| Cognitive Domain | Understanding of pain and cognitions. | Asking about or assessing pain related beliefs (including helpful/unhelpful beliefs, pain catastrophization etc) | 39 | Agree | Agree | 69.23% | No | Often |
|  |  | Asking the athlete what does this pain mean to them/ what is their understanding of their pain. | 39 | Agree | Strongly Agree | 87.18% | Yes | Often |
|  |  | Asking the athlete about their coping their strategies for pain. | 39 | Agree | Agree | 71.79% | Yes | Often |
|  |  | Asking the athlete about their pain related self-efficacy (confidence to manage their pain effectively). | 39 | Agree | Agree | 87.18% | Yes | Often |
|  |  | Asking about/assessing the level of attention the athletes pays to their pain. | 39 | Agree | Agree | 64.10% | No | Sometimes |
|  |  | Assessing cognitive traits/tendencies (explore/explicit, alertness/arousal). | 39 | Neutral | Somewhat Agree | 56.41% | No | Sometimes |
| Socioenvironmental Domain | Sports environment | Asking the athlete about pain culture within their sport/team. | 39 | Agree | Agree | 51.28% | No | Sometimes |
|  |  | Asking the athlete about the impact of the specific time of season on their pain. | 39 | Agree | Agree | 71.79% | Yes | Sometimes |
|  |  | Asking the athlete about pressure to perform/return to play for their team/sport. | 39 | Agree | Strongly Agree | 74.36% | Yes | Often |
|  | Support network | Asking the athlete about personal relationships and their support network. | 39 | Agree | Agree | 76.92% | Yes | Often |
|  |  | Asking the athlete about their home stress/commitments. | 39 | Agree | Agree | 82.05% | Yes | Often |
|  |  | Asking the athlete about their work/school/university relationships and support network. | 39 | Agree | Agree | 74.36% | Yes | Often |
|  |  | Asking the athlete about their work/school/university stress/commitments. | 39 | Agree | Agree | 76.92% | Yes | Often |
|  | Lifestyle factors | Asking the athlete about their alcohol or recreational drug intake. | 39 | Agree | Agree | 71.79% | Yes | Sometimes |
|  |  | Asking the athlete whether they are a current or previous smoker. | 39 | Agree | Agree | 58.97% | No | Sometimes |
|  |  | Asking the athlete about their general wellbeing/health. (Including physical, mental and social) | 39 | Agree | Agree | 87.18% | Yes | Often |
|  |  | Asking the athlete about/assessing their current quality of life. | 39 | Agree | Agree | 58.97% | No | Sometimes |
|  | Socioeconomic and sociocultural factors | Asking the athlete about the effect of pain/injury on their income and resources. | 39 | Agree | Agree | 71.79% | Yes | Sometimes |
|  |  | Asking the athlete about their access to appropriate sport and health facilities. | 39 | Agree | Agree | 79.49% | Yes | Often |
|  |  | Asking the athlete about their sociocultural background and pain culture. | 39 | Neutral | Somewhat Agree | 51.28% | No | Sometimes |
|  |  | Asking the athlete about their current level of education/employment. | 39 | Agree | Agree | 53.85% | No | Sometimes |
| General Aspects of Assessment | Aspects and considerations for athlete pain | Clinicians should be mindful not to over assess athletes when selecting pain assessment tools. | 38 | Agree | Strongly Agree | 86.84% | Yes | Often |
|  |  | Pain assessment tools should be selected based on the individual athlete and their pain timeframe. | 38 | Agree | Strongly Agree | 89.47% | Yes | Always |
|  |  | Time appropriate pain assessments (before/after training, on waking/at night etc.) at key time points agreed | 38 | Agree | Strongly Agree | 84.21% | Yes | Often |
|  |  | Knowing and understanding the athlete and their sport is an important aspect of athlete pain assessment. | 38 | Agree | Strongly Agree | 97.37% | Yes | Always |
|  |  | Developing a rapport and trust within the athlete-clinician relationship is an important aspect of the athlete | 38 | Agree | Strongly Agree | 97.37% | Yes | Always |
|  |  | Communicating clearly using health literate language so that the athlete understands the assessment is important to build confidence between the athlete and clinician. | 38 | Agree | Strongly Agree | 97.37% | Yes | Always |
|  |  | Clinicians should use the athlete pain assessment to educate, empower and guide the athlete. | 38 | Agree | Strongly Agree | 92.11% | Yes | Always |
|  |  | Clinicians should integrate technology into pain assessment data capture (wearables/mobile phone objective data capture and/or recording subjective updates) as appropriate | 38 | Agree | Agree | 55.26% | No | Sometimes |
|  |  | Clinicians should employ open and collaborative communication with the athlete's coach and wider network during the pain assessment process with the athletes permission. | 38 | Agree | Strongly Agree | 100.00% | Yes | Always |

## Discussion

This RTD provides a set of assessment items across six pain assessment domains. It is useful to consider why certain items met the threshold for consensus drawing on contemporary research and practical experience and consider the implications for research. Similarly, there is value in considering the reasoning behind the exclusion of certain aspects and comparing the value of a comprehensive pain assessment with the importance of practicality. Some limitations will also be considered.

### Neurophysiological Pain Assessment Domain

Neurophysiological assessment items such as pain location, quality and influencing factors are established aspects of a physiotherapist’s pain assessment.^13, 22, 23^ Thirteen items in the *pain characteristics, pain history* and *pain impact* categories achieved consensus to *always* be included as part of an assessment of upper and lower limb pain in athletes. Including the history of previous pain episodes is crucial for understanding the current pain presentation. Previous experiences and comparisons can significantly influence an athlete’s perception and impact of pain. This understanding can help guide the educational aspect of pain management.^5, 9^ The *impact of pain* category included assessment items related to both sports performance and daily activities, which are valuable indices of severity.^1^ Written measures such as pain diaries did not achieve consensus for inclusion, perhaps due to the time and effort to complete, despite the value of reflection and quantifying pain timelines during pain assessments.^21^

Pain severity scales are a staple of traditional pain assessment practice, however, the limitations of the available tools have been documented, including ceiling and floor effects, inability to compare athletes and pain episodes (reliability) and reducing the experience of pain to a single item.^12, 31^ Although previous research has highlighted how alternative pain scales may be more relevant to an athlete’s experience^40^ it was the Numerical Pain Rating Scale alone that met the threshold for inclusion, perhaps due to its ease and ubiquitous use in pain research and practice.^31, 52^ Consensus indicated a pain severity measure should be included *often*, not *always* perhaps indicating the limitations sports physiotherapists encountered in using these tools in practice. Alternative scales such as the traffic light system are embedded in clinical practice, particularly in the case of upper and lower limb tendon pain and should be considered in the next stage of developing the pain assessment framework.^48^ Understanding the dominant pain mechanism (nociceptive, neuropathic or nociplastic) is best practice in pain assessment both in general and athletic populations, guiding appropriate management.^22, 44^ However, formal measures of pain mechanisms did not meet the threshold for inclusion, in line with recent scoping review findings^42^ which found just one in four papers investigating pain assessment in athletes included a pain classification system, with those that did often choosing the outdated biomedical model. In line with previous qualitative research^40^, some sports physiotherapists commented how formal questionnaires and measures can be time-consuming, preferring to establish the pain mechanism using the pain interview questions such as asking about neurological signs and symptoms which participants felt should *always* be asked. Quantitative sensory testing did not achieve consensus for inclusion despite demonstrating value for quantifying pain sensitivity and predicting pain and function in research settings.^52^ The lack of perceived clinical utility aligns with previous findings that have demonstrated a lack of consensus regarding how they should be applied in practice.^18, 53^ Consensus on items regarding the physical aspects of assessment to identify the patient’s pain included *always* assessing range of motion and sports-specific functional tasks to identify the athlete’s specific pain. Special tests are a physical measure meeting consensus to be used *often*. Identifying the athlete’s specific pain experience is an important aspect of validating pain allowing the athlete to feel understood.^25, 35^

### Biomechanical Pain Assessment Domain

Biomechanical assessments include local measures of movement, strength and function as well as wider aspects like an athlete’s training load, sleep, nutrition, fatigue and fitness. Complimenting the physical aspects of neurophysiological pain assessment tools, sports physiotherapists felt testing full range of motion of local joints and assessing sport-specific postures and tasks while noting pain and symptoms were items that should *always* be included. These aspects align with current athlete pain assessment guidance and offer further opportunities to validate the athlete’s pain.^22, 35^ Measures of strength (such as repetition max, isokinetic dynamometry and local muscle endurance), balance (dynamic balance tests), motor control (quality of movement such as lumbopelvic or scapular control) and power (sprints/jumps) were aspects identified that should be included *often* to demonstrate the athlete’s movement profile. Pain, through arthrogenic muscle inhibition and other mechanisms, can impact performance and these markers provide a battery of tests to quantify impact in clinical practice.^33^ Fitness and agility and less challenging static balance and manual muscle testing (MMT) components were not included. Whilst MMT may be superseded by more objective measures it is frequently used in practice and remains a cornerstone of physiotherapy education, a useful option when more sophisticated and sport-specific measures are unavailable.^45^

Regarding wider biomechanical/biological considerations, the importance an athlete’s training load, occupational workloads and rehabilitation activities has been discussed widely including the IOC consensus statement on load.^50^ Sports physiotherapists opted to *always* include these measures indicating their relevance to the practical assessment of pain. The influence of wider lifestyle factors such as sleep, nutrition and menstrual cycle status on overall health and function, performance and pain perception has been established.^16, 24^ Sports physiotherapists indicated that they should *often* be included. Whilst biomarkers such as blood tests may form a part of diagnosis in sports and pain medicine this was not deemed to be necessary to include in a pain assessment framework.

### Affective Pain Assessment

The International Association for the Study of Pain definition of pain demonstrates that there is an emotional component that accompanies the sensory aspect of pain perception, highlighting the value of affective pain assessment strategies.^44^ Sports physiotherapists opted to include stress, mood and emotions such as fear, willingness to play through pain, adherence to rehabilitation and confidence to return to training/competition *often* as part of a pain assessment framework. The value of gauging an athlete’s understanding and tolerance for pain during rehabilitation and willingness to perform through pain has been established and may help guide clinical decision-making.^3, 49^ Understanding feelings and emotions may improve an athlete’s interoceptive capabilities which can have positive effects on sporting performance and general health.^47^ Additionally, screening for specific mental health diagnoses is something sports physiotherapists felt is *sometimes* required. Qualitative research findings have highlighted the emotional toll of pain and the lack of assessments in this area with a strong athlete voice calling for their inclusion, indicating the importance of these aspects.^10, 39–41, 56^ Understanding or assessing specific personality traits did not meet consensus.

### Cognitive Pain Assessment

The cognitive aspects of assessment provide a window into the athlete’s understanding of their pain and the internal context of their pain experience.^23^ Asking about or assessing pain-related beliefs using the validated Athlete Fear-avoidance Questionnaire and the Pain Catastrophising Scale narrowly missed consensus reflecting their lack of use in pain research and practice.^42^ Given the importance of beliefs in the perception and experience of pain^3, 5^ and how they inform an athlete’s actions and the management of their pain^8, 9, 52^ this is something that should be reconsidered when further developing the pain assessment framework. In contrast, asking an athlete “What does your pain mean to you/What is your understanding of your pain?”, alongside asking about coping strategies and their level of confidence (efficacy) to manage their pain all achieved consensus to be included *often*. These items are less time-demanding and translate to practice and the pain management process.. The cognitive pain assessment domain is the least frequently used in both research and practice.^42^ Including the items highlighted here by sports physiotherapists may help fill this void. Distraction and managing attention and focus are tools sometimes employed by athletes during the pain management process.^10, 39^ Both athletes and clinicians have expressed concerns about the overassessment of pain increasing the athlete’s attention on and therefore amplifying, their pain.^40, 56^ Despite the potential value of exploring attention and distraction, they did not achieve consensus, with the potential for implicit assessment of these aspects through the initial interview.

### Socioenvironmental Pain Assessment

The impact of context on injury prevention and pain management has been established^7, 10, 52^ including in athlete pain guidelines.^22^ Whilst an athlete’s internal context can be best understood by considering affective and cognitive factors, the external context of athlete pain experience can be optimally expressed and assessed by considering the socioenvironmental aspects of pain.^22^ Of note, asking athletes about the pain culture within their sport or team did not meet consensus. Perhaps an appreciation of the prevailing culture is something that is gauged implicitly through a wider understanding of the team/sport. Indeed, previous qualitative insights have outlined how an attitude of respect and response to pain rather than ignoring pain may positively influence an athlete’s pain perception.^3, 5, 10, 39, 56^

Understanding the time of the season (s*ometimes*) and pressure to perform or return to play an athlete feels within their specific sports environment (*often*) were deemed to be key aspects for inclusion.

Regarding lifestyle factors, asking about general well-being met the threshold to be included *often*, with asking about alcohol/recreational drugs being deemed necessary *sometimes,* likely when suspected to be a contributor in specific presentations. Quality of life was not included with some participants commenting this is not something they ask but gauge implicitly throughout the assessment. The effect of pain on income/resources (*sometimes*) and access to appropriate facilities and healthcare (*often*) were, with education/employment and sociocultural background and pain culture not meeting the threshold. Although education and sociocultural aspects have been highlighted as key prognostic indicators in the literature when it comes to overall health and development of chronic pain,^32, 55^ sports physiotherapists tended to favour questions related to modifiable aspects that have practical solutions and implications. Although this consensus may facilitate the development of a practical and implementable framework it is important not to lose sight of the wider aspects influencing an athlete’s pain that are significant in understanding their unique presentation.

Highlighting the wider socioeconomic and sociocultural aspects may help facilitate systemic changes that may be needed in the future (e.g., social policy).^55^

### General Aspects of Pain Assessment

This domain identifies key components of the assessment process and conduct that accompany practical assessment items. The nine items in this domain were included based on recent evidence surrounding communication, timing, knowledge/education, relationships and the integration of technology including clinical guidelines^4, 30^ and qualitative insights.^10, 11, 40, 41^ Items based on effective and clear communication, building a rapport/trust and getting to know the athlete and their sport as well as using the assessment as an opportunity to educate the athlete about their pain all met the consensus to *always* be included. Communicating and collaborating with the athlete’s support network and coaches, with the athlete’s permission, achieved 100% consensus and was deemed something that should *always* be considered, highlighting the role of communication in athlete welfare and performance aligning with findings from the literature.^11, 27^ Furthermore, the importance of clear communication, considering principles of health literacy and using plain uncomplicated language is an aspect sports physiotherapists felt should *alw*ays be included. Clear communication has been linked to positive outcomes in healthcare and sport, particularly when assessing and explaining pain and injury.^6, 27, 32^ Sports physiotherapists opted to include knowing and understanding the athlete and their sport, in essence, their internal (pain perception and experiences) and external (sports environment) context as something that is *always* required, underscoring the importance of context.^7, 52^ Pain neuroscience education or ’explaining pain’, has been established as an important aspect of pain management in athletes.^38^ Effective communication and explanation of pain during the assessment guides the management process with one athlete in our previous research^40^ stating;

*“I know exactly what it is, and I nearly come to terms with it an awful lot quicker.. I feel like my education around it has a huge impact..”.* – A08

Additionally, considering the individual and their pain timeline when selecting pain assessment items was deemed something that is a*lways* required. This links with the need to be judicious when selecting pain assessment items, avoiding over-assessment and selecting appropriate pain assessment items at key time points agreed with the athlete (e.g. before/after training/competition) which sports physiotherapists felt is *often* required and facilitates the clinical reasoning process prioritised in clinical practice guidelines.^30^ These three aspects taken together speak to the need for an individualised and selective pain assessment that requires clinical reasoning from the sports physiotherapist to understand pain mechanisms, tissue healing and pain timelines (acute, subacute and chronic) and the spectrum from sports injury to sports-related pain^25^ and may ultimately ease the assessment burden on both athletes and physiotherapists who operate in the time-pressured sports performance environment.^6^

Finally, the integration of technology into the assessment did not meet consensus despite its value in healthcare and performance and the integration within national physiotherapy frameworks^34^ and the ubiquitous role of technology in the life of the modern athlete. This may be due to a reticence on the physiotherapist’s part to add additional inputs to over encumbered athletes with collection of data that may be irrelevant to the pain presentation. Some clinicians commented they were wary of adding additional smartphone use to athletes who are inundated with information. Additionally, currently available technologies that may not have been developed in partnership with physiotherapists and athletes may fail to meet the specific demands of assessments in sport, with guidelines highlighting the importance of co-creation and user-centred design in technology and connected health.^34^

### Limitations and Considerations for Framework Development

On one hand, the development of a practical pain assessment framework necessitates the inclusion of a set of measures that can be integrated into the clinical workflow of a sports physiotherapist working across a variety of clinical settings and at times with suboptimal facilities.^6^ Consensus for cognitive, affective and socioenvironmental (contextual) factors in this study often favoured these practical approaches that may inform pain management. On the other hand, prioritising a comprehensive assessment that appreciates and represents the athlete’s pain experience encourages physiotherapists to challenge their current practice to consider alternative methods and measures to ensure the wider contextual is understood. Whilst some outcome measures that address these wider aspects may be burdensome, or their inclusion may seem arbitrary when information related to an aspect of pain assessment might be gathered implicitly, it is important to recognise the use of standardisation and protocols in optimising health and performance outcomes and reducing clinical errors and omissions, particularly in a realm where athletes often feel misunderstood. Future work should focus on distilling the findings of this RTD into a practical yet comprehensive pain assessment tool that clinicians and researchers can use.

Although there was a high completion rate (98%), assessment items in the latter portion of the survey were completed by less participants than those in the earlier portion. Additionally, the number of consensus reviews differed across the six assessment domains. Including randomisation of survey domains and a mandatory review of consensus in all domains should be considered in future RTD studies.

## Conclusion

This RTD study with international sports physiotherapists presents a comprehensive set of recommendations for assessing upper and lower limb pain in athletes. Our findings highlight the importance of a multidimensional approach, incorporating neurophysiological, biomechanical, affective, cognitive, socioenvironmental, and general assessment aspects. The consensus-derived recommendations offer practical, implementable strategies for routine clinical use, balancing the need for thorough assessment with the practical constraints of sports physiotherapy practice.

It is hoped that these recommendations can guide clinicians in delivering more comprehensive and context-sensitive pain assessments, ultimately improving pain management and rehabilitation outcomes for athletes. Future work should focus on refining these recommendations into a cohesive, user-friendly framework that can be validated and integrated into clinical settings.

## Practical Implications

### Findings

- We present a comprehensive and practical set of expert sports physiotherapist consensus-derived pain assessment items across six key domains for pain assessment. (20 neurophysiological, 15 biomechanical, 8 affective, 3 cognitive, 10 socioenvironmental and 8 assessment aspects) alongside the frequency with which they are required. (29 *always*, 32 *often*, 4 s*ometimes*).

### Implications

- This study establishes expert consensus regarding the choice and frequency of pain assessment items for athlete pain across six domains: neurophysiological, biomechanical, affective, cognitive, socioenvironmental and general aspects of assessment. Sports physiotherapists can use these recommendations to inform their pain assessment with athletes experiencing upper and lower limb pain.

### Caution

- Consideration should be given to aspects that may have been excluded due to perceived practical feasibility to ensure a comprehensive assessment is delivered with clinicians being encouraged to apply clinical reasoning and judgement in each athlete scenario.

## Study Details

### Institutional Review Board Approval

Ethical permission was granted for our study by the Univerities Human Research Ethics Committee. (LS-22-40-xxx-xxx)

## Acknowledgements

We would like to thank Prof Kieran O’Sullivan for providing expert consultation regarding the development of the initial Delphi Survey.

## Author Contribution Statement

CP conceived the original idea and developed the initial survey. CP, BC, BF and TW developed the original idea and survey. CP completed data collection and data analysis. BC, BF and TW reviewed the data analysis. CP composed the initial manuscript draft. BC TW and BF provided comments on and contributed towards the writing and editing of the final draft. All authors have read and agreed to the published version of the manuscript.

## Patient, athletes and public involvement statement

A PPI panel of athletes as described in the methods section was integral in the conduct of this research project. The panel have been involved in the previous aspects of this research which involved focus groups of athletes and physiotherapists and provided feedback towards the development of this paper. The PPI panel will continue to be involved with the final development of the pain assessment framework, the next stage of this research project.

## Data Sharing

Data are available through the Open Science Framework (OSF) which is a public open access repository and can be accessed at 10.17605/OSF.IO/D8T3N

