## Supplementary material for "Developing Consensus for an Upper and Lower Limb Athlete Pain Assessment Framework – A Real-time Delphi Study with International Sports Physiotherapists": Figure 1

**Funding:** This work was supported by funding from Science Foundation Ireland under the grant for the Insight SFI Research Centre for Data Analytics (SFI/12/RC/2289_P2) Funders had no role in the data collection, analysis or interpretation and will have no role in approving the final manuscript.

**Financial disclosure and conflict of interest:** I affirm that I have no financial affiliation (including research funding) or involvement with any commercial organization that has a direct financial interest in any matter included in this manuscript, except as disclosed and cited in the manuscript. Any other conﬂict of interest (i.e., personal associations or involvement as a director, officer, or expert witness) is also disclosed and cited in the manuscript.

**Institutional Review Board** Ethical permission was granted for our study by the UCD Human Research Ethics Committee. (LS-22-40-Purcell-Caulfield)

**Word Count:** 4432/4500

**Acknowledgements**: We would like to thank Prof Kieran O’Sullivan for providing expert consultation regarding the development of the initial Delphi Survey.

**Author Contribution Statement**	CP conceived the original idea and developed the initial survey. CP, BC, BF and TW developed the original idea and survey. CP completed data collection and data analysis. BC, BF and TW reviewed the data analysis. CP composed the initial manuscript draft. BC TW and BF provided comments on and contributed towards the writing and editing of the final draft. All authors have read and agreed to the published version of the manuscript.

**Patient, athletes and public involvement statement.**

A PPI panel of athletes as described in the methods section was integral in the conduct of this research project. The panel have been involved in the previous aspects of this research which involved focus groups of athletes and physiotherapists and provided feedback towards the development of this paper. The PPI panel will continue to be involved with the final development of the pain assessment framework, the next stage of this research project.

**Data Sharing**

Data are available through the Open Science Framework (OSF) which is a public open access repository and can be accessed at 10.17605/OSF.IO/D8T3N
