## Appendices for "Developing Consensus for an Upper and Lower Limb Athlete Pain Assessment Framework – A Real-time Delphi Study with International Sports Physiotherapists"

### **Appendix A – Real-time Delphi initial Survey**

**Overview provided for PPI panel and pain expert reviewers**

- This survey will present a range of tools and concepts that **could** be included in a pain assessment framework for Physiotherapists working with athletes with upper and lower limb pain. All items that receive **70%** consensus or greater will be taken forward and included in a practical framework/ clinical workflow that Physiotherapists can use **as an adjunct** when completing an assessment.
- The pain assessment framework that is developed from this study will serve as a memory aid or toolkit that can be consulted to ensure physiotherapists get the most out of their assessment. The framework will not be a strict recipe for clinicians to follow, but rather a flexible guide that reflects the unique nature of each assessment, athlete preferences and the role of the physiotherapist’s clinical reasoning and experience.
- Items are divided into 6 sections. Sections 1-5 correspond to the 5 International Olympic Consensus statement Athlete Pain domains (neurophysiological, biomechanical, affective, cognitive, and socioenvironmental), and a separate section which includes aspects of assessment that should be considered. (Hainline, 2017)
- The items included in the survey have been chosen based on the results of a recent scoping review and focus groups of athletes and sports physiotherapists completed by the research team.

**Text to appear at the start of all sections;**

***“I feel this pain assessment item is important when assessing upper and lower limb pain in athletes and should be included in the pain assessment framework:***

**6-point Likert scale (Select one)**

Strongly Disagree – Disagree – Somewhat Disagree– Somewhat Agree – Agree – Strongly Agree

The comments box below is available to provide rationale/ thoughts related to your vote

_____________________________________________________________________________________________________

***“How often do you feel this pain assessment is/will be required in practice.”***

**5-point Likert scale (Select one)**

Never– Rarely – Sometimes –Often – Always

The comments box below is available to provide rationale/ thoughts related to your vote

______________________________________________________________________________________________________

**Section 1. Neurophysiological Pain Assessment Measures.**

***Pain Characteristics***

Subjective questions about the various characteristics of pain.

1. Locating the specific site of the athlete’s pain through questioning (e.g. can you pinpoint your pain?)
2. Asking the athlete about the nature of their pain which may include specific adjectives or descriptors.
3. Asking about the irritability of the athlete's pain (how long it takes to come on/go away).
4. Asking about night pain.
5. Asking about the 24 hour pattern of pain.
6. Asking about aggravating and easing factors.
7. Asking the athlete about pain medication (current, previous, or recent changes).

***Pain History***

Subjective questions to establish the timeline, history and context of an athlete's pain.

8. Asking about the athlete’s specific pain history and experiences and establishing the context of this pain presentation.
9. Asking about pain duration (e.g. acute, subacute, chronic).
10. Asking about the onset and timeline of pain. (e.g. gradual onset, sudden onset, recurrent, etc).
11. Asking the athlete to compete a pain diary and/or reflections in their own time.

***Impact of Pain***

Subjective questions about the impact of pain on sport and everyday life.

12. Asking the athlete about the impact of their pain on their activities of daily living.
13. Asking the athlete about the impact of their pain on their sports performance. (e.g training and/or competition).
14. Asking the athlete about specific sporting tasks that pain is experienced during/ exacerbated by.

***Pain Scales and Classification***

Scales to capture the intensity of pain and measures to classify pain mechanisms

15. The Numerical Pain Rating Scale (NPRS) also known as the 0-10 pain scale.
16. The Visual Analogue Scale (VAS).
17. The Pain Faces Scale. (Could be used with Paediatric athletes).
18. A Descriptive pain scale e.g. (mild, moderate and severe or threatening non-threatening).
19. *Establishing the underlying pain mechanism based on the International Association for the Study of Pain (IASP) pain mechanisms. classification through questionnaires/checklist (e.g, LANSS, S-LANSS, NPQ, DN4 or PainDETECT for neuropathic pain, Smart et al., 2011 Mechanisms based MSK pain checklists).

LANSS = Leeds Assessment of Neuropathic Symptoms and Signs S-LANSS = Self-complete Leeds Assessment of Neuropathic Symptoms and Signs NPQ = Neuropathic Pain Questionnaire DN4 = Douleur neuropathique en 4 questions

***Neurological Aspects***

Assessment tools and strategies related to neurological screening and neurodynamic assessment of the upper and lower limb.

20. Asking about neurological signs and symptoms. ( shooting/stabbing type pain, loss of sensation, loss of power, pins and needles and other paraesthesia).
21. Completing a neurological examination (myotomes/dermatomes/reflexes).
22. Assessing neurodynamics (through upper limb neural tension tests SLUMP/ Straight leg raise etc).
23. Screen for neuropathic pain using validated measures (eg. PainDETECT, DN4).

***Physical or Manual Pain Assessment***

Physical, manual or ''hands-on'' assessment tools and techniques to locate, identify, provoke or characterize an athlete‘s pain.

24. Completing Quantitative Sensory Testing (e.g. pressure pain threshold, pain tolerance, hot/cold sensitivity.)
25. Locating the specific site of the athlete’s pain through palpation.
26. Pain provocation/identification of the athlete’s pain through range of motion.
27. Pain provocation/identification of the athlete’s pain using Special Tests (e.g. Empty Can/Hawkins Kennedy for shoulder, McMurray's test for knee, talar tilt for ankle.
28. Pain provocation/identification of pain through athlete-identified functional tasks. (E.g climbing/descending stairs, brushing hair, carrying bags etc.)

**Section 2. Biomechanical Pain Assessment Measures.**

***Movement Related tools and tests***

Tools including strength, range of motion, balance, motor control, power, agility and fitness assessments.

1. Objective strength assessments (handheld dynamometry, isokinetic strength testing, repetitions max testing etc).
2. Local muscle endurance tests (e.g. single leg hamstring bridge/ single leg squat to box to failure).
3. Manual muscle Testing (Oxford Grading etc).
4. Passive and active range of motion testing (noting quality and quantity of available range +/- pain/stiffness).
5. Static balance measures (Eyes Open/Eyes Closed, single leg, tandem, unstable surface Romberg etc).
6. Dynamic balance measures (Y-Balance Test, Star Excursion Balance Test, multi-directional hopping.
7. Motor control (landing mechanics, lumbopelvic control, trunk control, scapular control etc).
8. Sport-specific movement or posture assessment (eg Squat, lunge, running, jumping, throwing, kicking).
9. Power/speed assessment (timed sprint, CMJ, broad jump, single leg hop, triple hop, reactive strength index etc).
10. Agility Test ( 5-0-5, Illinois, t-test etc).
11. Fitness Test (V02 max or submaximal testing, bleep test, intermittent shuttle test, timed test to exhaustion, bronco etc).

***Wider Biological Tools and Tests***

Tools including acute and chronic training load, current activities and rehabilitation, fatigue, sleep, nutrition, menstrual status and blood markers.

12. Establishing current training load.
13. Establishing current *rehab/prehabilitation activities.
14. Establishing chronic training load/ capacity.
15. Asking the athlete about current level of general and sports related fatigue/tiredness (or asking about unusual or increased levels of fatigue).
16. Asking about sleep duration and quality.
17. Asking about nutrition quality/status or whether the athlete is meeting current energy demands.
18. Asking females about their menstrual cycle status/regularity and the influence on pain.
19. Asking the athlete about results from recent blood tests, biomarkers etc.
20. Asking athletes who work about occupational-
21. related loads

**Section 3. Affective Pain Assessment Measures.**

***Affective/psychological pain assessment measures***

Subjective questions related to mood, general wellness, stress mental health, psychological readiness to compete, training through pain and various personality traits.

1. Asking about mood, feelings and emotions related to pain and/or sport.
2. Asking about/assessing general stress.
3. Asking about/assessing sports-related stress.
4. Asking about past or present mental health or learning disorder diagnoses (eg anxiety, depression, bipolar, eating disorders, attention deficit hyperactivity disorder, dyslexia etc).
5. Asking about psychological readiness/confidence to train/compete.
6. Asking about willingness to exercise/train/ with pain.
7. Asking about/assessing risk-taking versus safety personality traits.
8. Asking about/assessing rehabilitation adherence/motivation.

**Section 4. Cognitive Pain Assessment Measures.**

***Tools and questions related to the understanding of pain and cognitive aspects***

Asking about or assessing pain beliefs, thought processes and patterns, self-efficacy, coping strategies and cognitive traits.

1. Asking about/ assessing pain-related beliefs (including helpful/unhelpful beliefs, pain catastrophization etc).
2. Asking the athlete what does this pain mean to them/ what is their understanding of their pain.
3. Asking the athlete about their coping strategies for their pain.
4. Asking the athlete about their pain-related self-efficacy (confidence to manage their pain effectively)
5. Asking about/assessing the level of attention the athlete pays to their pain
6. Assessing cognitive traits/tendencies (explore/exploit, alertness/arousal)

***Section 5. Socioenvironmental Pain Assessment Measures.***

***Sports environment***

Elements such as pain culture, time of season and pressure to return to play.

1. Asking the athlete about pain culture within their sport/ team.
2. Asking the athlete about the impact of the time of season on pain
3. Asking the athlete about the pressure to perform/return to play for a team/sport

***Support Network***

Asking the athlete about their relationships, stress and commitments at home and in work or school/university.

4. Asking the athlete about personal relationships and support network

(this includes close family, partner, spouse etc and the support and/or effect that difficult or challenging family situations/life events may have on pain)

5. Asking the athlete about home stress/commitments.
6. Asking the athlete about work/school/university relationships and support networks. (this includes friends, peers, colleagues, mentors etc and the support and/or the effect that difficult or challenging circumstances within these relationships may have.)
7. Asking the athlete about work/school/university stress/commitments.

***Lifestyle Factors***

Asking about alcohol, smoking, general well-being and health and assessing quality of life.

8. Asking the athlete about alcohol intake
9. Asking the athlete whether they are a current or previous smoker
10. Asking the athlete about general wellbeing/health. (Including physical, mental and social)
11. Asking the athlete about/assessing current quality of life

**Sleep, Nutrition, Menstrual status etc. are categorized as Wider biological pain assessment tools as part of the IOC Athlete Pain Framework (2017)*

****Socioeconomic and sociocultural factors***

Asking about wider social, economical and cultural aspects and access to resources.

12. Asking the athlete about the effect of pain/injury on income and resources (e.g. access to healthcare may be affected if athletes dropped from the team etc)
13. Asking the athlete about their access to appropriate sport and health facilities.
14. Asking the athlete about sociocultural background and pain culture
15. Asking the athlete about the current level of education/employment

**Section 6. Aspects of Pain Assessment**

***Aspects and considerations for athlete pain assessments.***

***Aspects relating to the structure of assessment, communication considerations, means of collecting pain information and developing the athlete-physiotherapist relationship.***

1. Clinicians should be mindful not to over-assess athletes when selecting pain assessment tools.
2. Pain assessment tools should be selected based on the individual athlete, their diagnosis and their pain timeframe.
3. Athletes should record their own pain at key time points identified by themselves and agreed with their clinician. (before/after training, on waking/at night etc.)
4. Knowing and understanding the athlete and their sport is an important aspect of athlete pain assessment.
5. Developing a rapport and trust within the athlete-clinician relationship is an important aspect of the athlete pain assessment
6. Communicating clearly using health-literate language so that the athlete understands the assessment is important to build confidence between the athlete and clinician.
7. Clinicians should use the athlete pain assessment to educate, empower and guide the athlete
8. Clinicians should integrate technology into pain assessment data capture (wearables/mobile phone objective data capture and/or recording subjective updates) as appropriate.
9. Clinicians should employ open and collaborative communication with the athlete‘s coach and wider network during the pain assessment process with the athlete’s permission.

### **Appendix B – Images of Surveylet (Calibrum) real-time delphi software.**

Image A – Displays a the anonymous real-time delphi survey format participants were with.

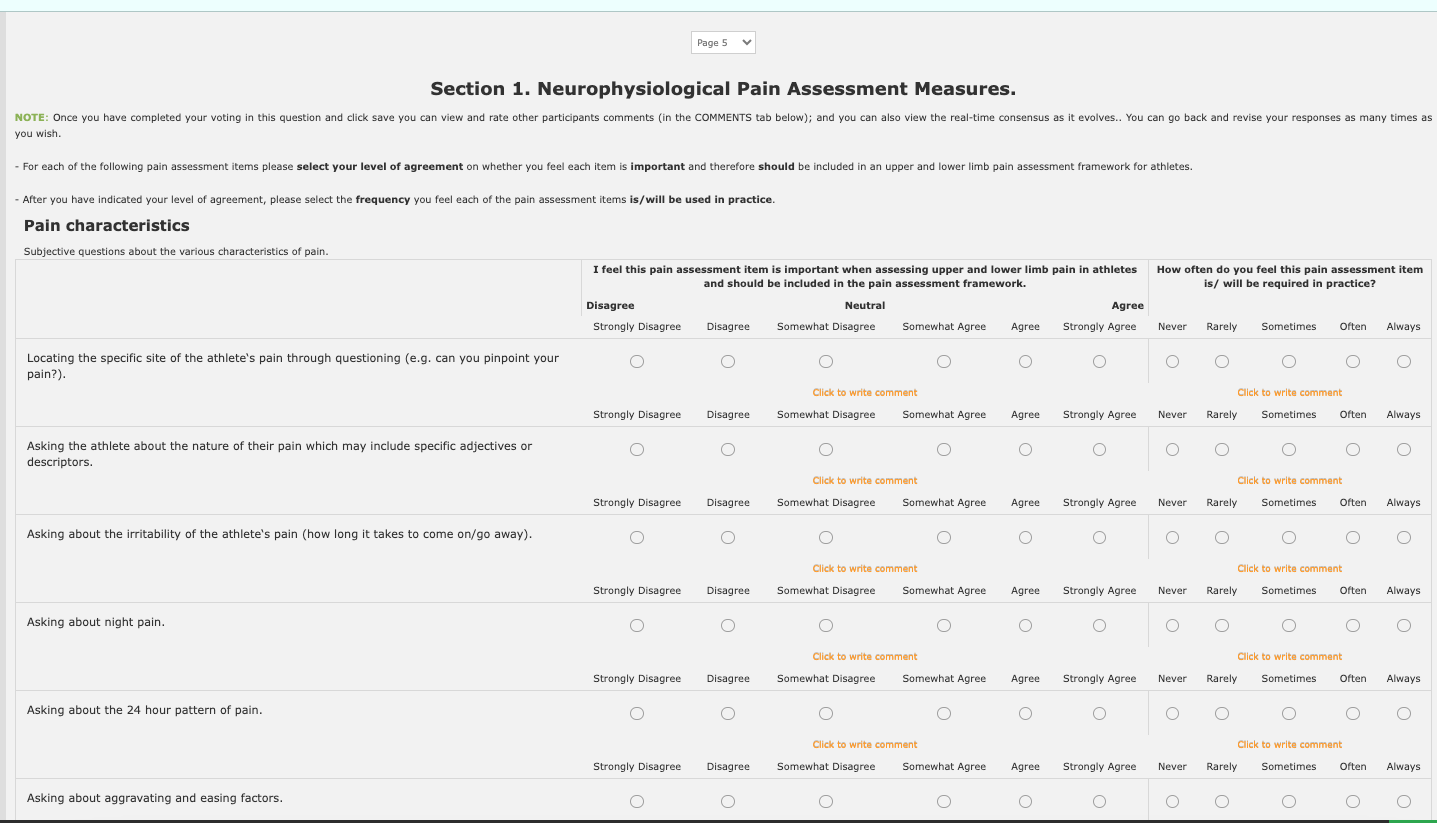

Image B – Displays the anonymous consensus graphs and comments participants were provided with following the completion of each section of the real-time delphi survey.

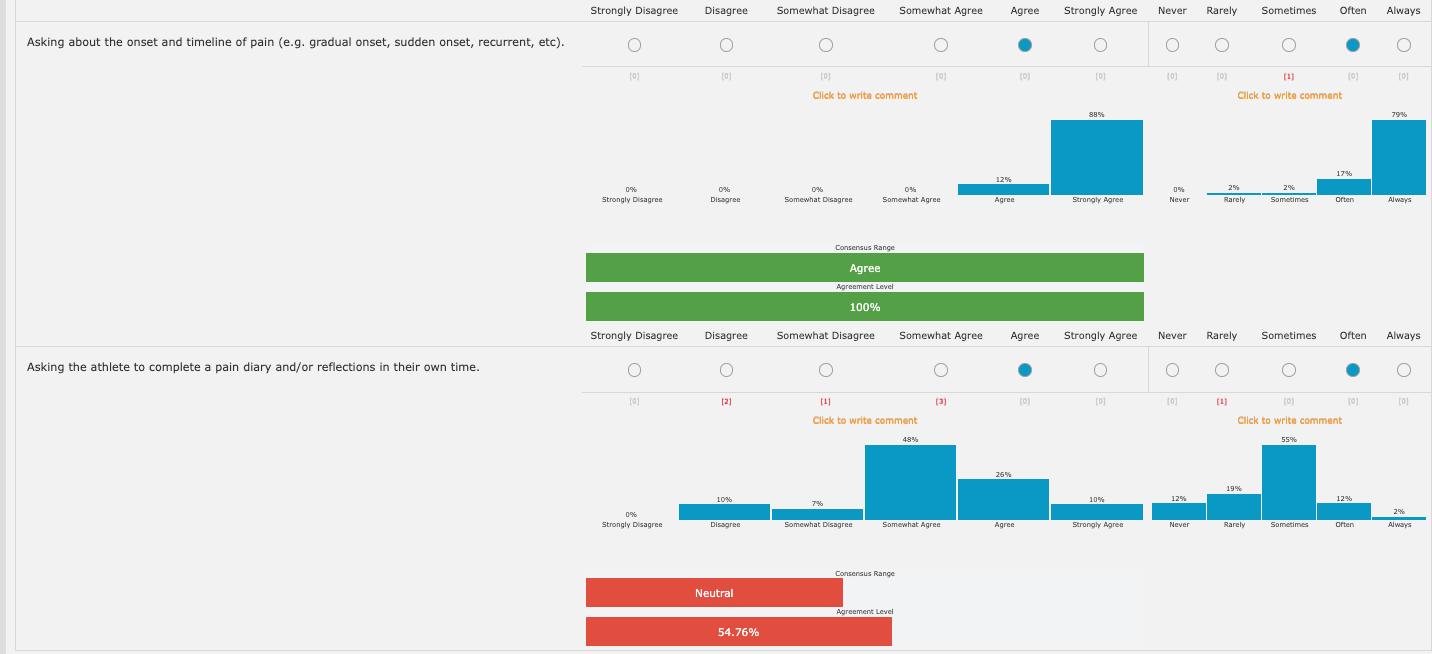
